# A retrospective analysis of climate-based dengue virus transmission suitability and demographic changes over the last four decades

**DOI:** 10.1101/2023.11.21.23298791

**Authors:** Taishi Nakase, Marta Giovanetti, Uri Obolski, José Lourenço

## Abstract

The geographical range and intensity of dengue virus transmission have significantly increased in recent years due to changes in climate, urbanization and human movement. Using estimates of dengue transmission suitability based on historical climate data, we analyze the effects of climate and demographic changes on the global population living in high-risk areas over the last four decades. We find that climate-related stress has been greatest in the Global South, especially in Africa and Southeast Asia. Although the geographic limits of dengue transmission suitability remained relatively stable in recent years, the global population at risk within those areas has grown by approximately 1.7 billion, driven by population growth in areas with historically dengue-favorable climate conditions. While many studies focus on future changes, we provide oft-overlooked evidence towards understanding how past climate and demographic change may have shaped the current global epidemiology of dengue.

**Teaser:** Retrospective analysis of climate and demographic changes reveals growth in global population in high-risk DENV settings.

## Introduction

Dengue is the most common mosquito-borne viral disease globally, with an estimated 60 million symptomatic infections per year across 130 countries (*1*, *2*). The dengue virus (DENV) is primarily transmitted by *Aedes aegypti* mosquitoes, which typically inhabit urban environments in tropical and subtropical regions. Disease burden in affected countries has greatly increased over the past few decades with Southeast Asia, South America, and the Western Pacific accounting for the majority of reported cases (*1*). In the last decade, the geographical range of dengue has also expanded to more temperate climates with the establishment of epidemic activity in parts of North America and a rise in autochthonous transmission in southern Europe (*2*, *3*). These recent trends are thought to be driven by changes in climate, land use, urbanization and human movement that increase proximity between vectors and humans and favor the spread of vector species (*4*). While improvements have been made in the ability to estimate the global distribution of current and future dengue risk (*5–7*), the extent to which environmental changes have influenced the geographical limits of DENV transmission in past decades has not yet been thoroughly examined. Given the significant health and economic threats posed by DENV, understanding the role some of these factors may have played in recent shifts in DENV epidemiology is essential for effective health systems planning.

The geographical range, timing and intensity of dengue transmission are influenced by the interaction between environmental and ecological factors including temperature (*8*, *9*), humidity (*10*), urbanization (*11*, *12*), human mobility (*12*) and the presence of water sources (*13*). Climate strongly influences dengue transmission because it modulates several physical and behavioral traits of mosquitoes such as adult female lifespan, aquatic developmental rates, viral incubation period and biting rate, thereby altering a mosquito’s physiological potential to transmit the virus to new hosts (*9*, *14*). In experimental studies of *Ae. aegypti* mosquitoes, higher temperature increased transmission potential through, for example, increases in mosquito lifespan (*15*) and decreases in viral incubation period (*14*). At extreme temperatures, however, some mosquito traits such as survival are adversely affected (*9*). At the same time, higher relative humidity has been shown to increase mosquito lifespan (*16*) and enhance virus propagation (*17*) in several settings. Using accumulated experimental data on *Ae. aegypti* mosquitoes, it is now possible to mechanistically model these relationships between climate variables and mosquito-viral traits (*18*).

In recent years, several studies have used experimental data to inform and parametrize DENV suitability measures that model and quantify the interactions between climate and mosquito-viral traits (*15*, *18–21*). Suitability measures vary in methodology and interpretation, but virtually all proposed approaches aim at quantifying transmission potential, often without resorting to epidemiological data (i.e. reported cases). Since climate modulates the spatiotemporal dynamics of DENV transmission, these mechanistic models are useful in defining the geographical limits and seasonal patterns of transmission across both endemic and emerging areas. In contrast, statistical approaches that estimate associations between climate variables and dengue cases are restricted in their ability to define the changing geographical limits of DENV transmission (*5*, *6*). This is due to a combination of sparse reporting in areas where transmission is low and difficulty capturing the complex nonlinear interactions between variables that mechanistically drive transmission. Conversely, prior mechanistic modeling approaches have based their estimates of dengue transmission solely on temperature without consideration of the influence of other climate variables (*15*). Furthermore, many mechanistic models have been developed on locally specific parameter estimates with applications limited over small spatial and temporal scales (*22–24*).

In this study we quantify spatiotemporal trends in climate-based dengue transmission suitability at a high spatial resolution over the last four decades using a widely validated mechanistic DENV suitability measure for *Ae. aegypti* mosquitoes referred to as Index P (*19*, *24–26*). Index P is a proxy for the transmission potential of a single adult female mosquito under conditions where susceptible hosts, the virus and its vectors are assumed to be present. We use a large collection of spatiotemporal maps of Index P for 186 different countries and territories based on satellite temperature and relative humidity time series from 1979 to 2022. This data has been recently validated using high-resolution dengue incidence data from Brazil and Thailand, providing robust estimates of long-term, climate-based DENV suitability trends (*26*). Here, we quantify climate-based changes in DENV transmission suitability globally and identify where and when climate conditions were likely to be favorable to transmission. The identified long-term trends in climate-based DENV transmission suitability are then analyzed against global demographic changes to understand how the distribution and size of human populations living in areas with high suitability have evolved over time. This approach highlights geographical heterogeneity in the evolving climate-based limits of DENV transmission and offers deeper insight into the synergistic roles climate and demographic changes may have played in the global increase in dengue epidemic activity observed over the last several decades.

## Results

### Summarizing climate-based suitability for DENV transmission and persistence

Transmission suitability is the raw quantification of transmission potential per adult female mosquito, which we summarized by calculating the average transmission suitability for 1979- 2022 (**Fig. 1A, Fig. S1-S2**). It highlights areas that remain suitable for transmission throughout the year and also accounts for areas that are characterized by brief periods of very high suitability interspersed with periods of less favorable climate conditions. It should thus be interpreted as the potential for epidemic activity rather than endemic transmission.

**Fig. 1.**
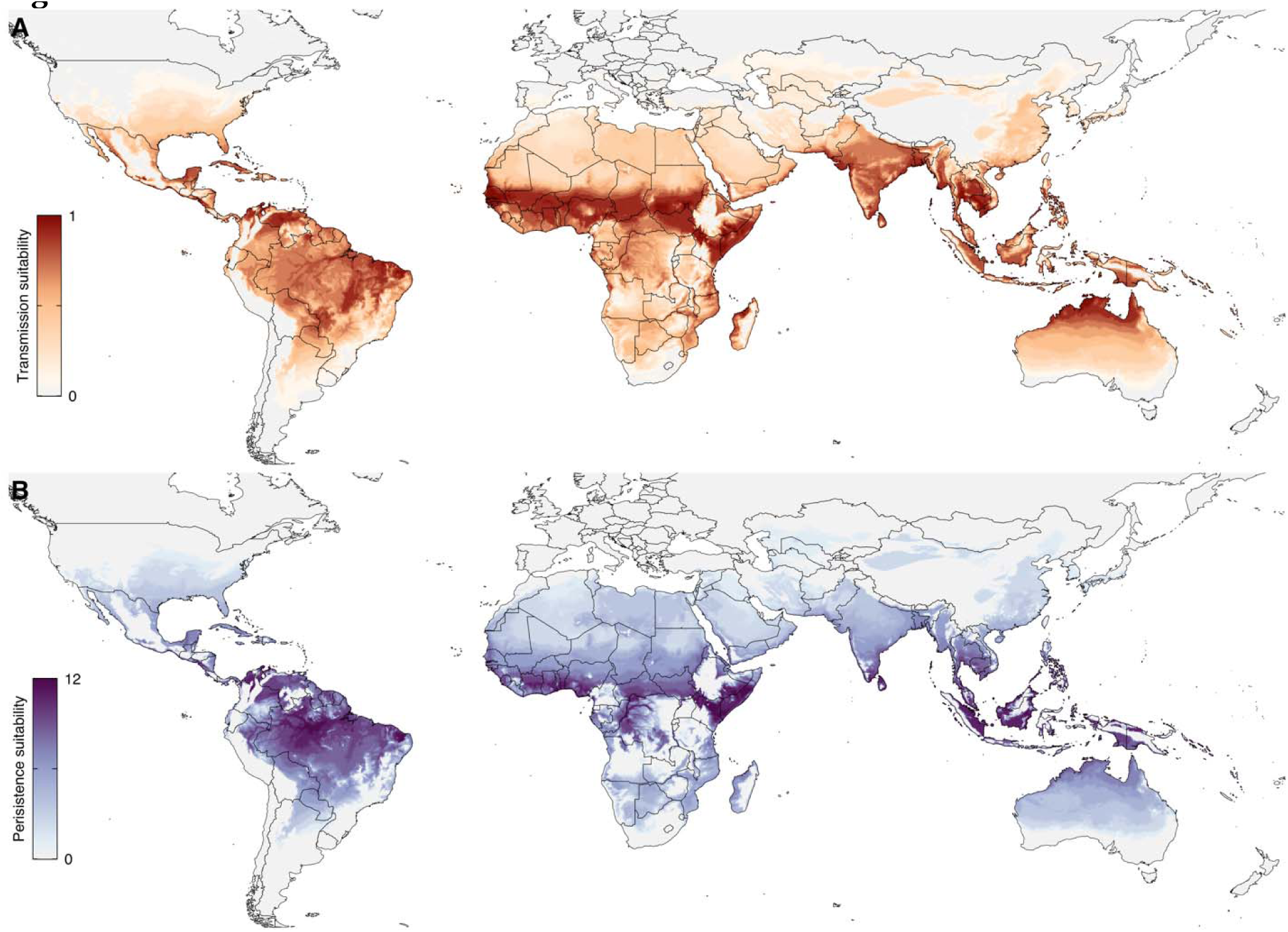
Global map of climate suitability for DENV transmission and persistence for 1979-2022. (**A**) For visualization purposes, average transmission suitability is presented on a logarithmic scale (normalized by maximum value). Transmission suitability is calculated by averaging the transmission potential over the selected time interval. (**B**) Persistence suitability is measured as the number of months the transmission potential exceeds a threshold of 1.0 over a year, averaged over the selected time interval.

High levels of transmission suitability were estimated for many regions within the tropical, subtropical, and temperate zones (also summarized globally in **Fig. S3** for each of the 28 Köppen-Geiger climate types). The spatial distribution of transmission suitability was largely consistent with reported dengue cases across the western tropical zone of Africa, the Indian subcontinent, Southeast Asia, the Western Pacific and South America (*2*). High transmission suitability was also estimated in areas where DENV has shown autochthonous transmission but is not yet known to be endemic (e.g. southern United States and northern Australia), or where infections are suspected to be under-reported due to limited surveillance (e.g. Central and Eastern Africa). Transmission suitability was relatively low across continental Europe with areas of non-zero suitability confined to isolated parts of Spain, France, Italy, and Turkey.

Persistence suitability is the number of months in a year that suitability exceeds a threshold deemed favorable for DENV transmission. It was summarized by calculating the average number of months with high transmission suitability per year for 1979-2022 (using a suitability threshold of 1.0) and can be interpreted as a proxy of the potential for endemic transmission (**Fig. 1B, Fig. S4**).

The spatial distribution of transmission suitability (**Fig. 1A**) was generally consistent with that of persistence suitability (**Fig. 1B**), with the overlapping peak suitability around the equatorial tropical region. There were, however, some notable differences between the transmission and persistence suitability maps, indicating local restrictions to endemicity. Relatively few areas had high year-round transmission suitability. These areas included parts of Southeast Asia, Eastern and Western Africa and northern Brazil, where DENV is known to be largely endemic. Many areas with high transmission suitability had relatively low persistence suitability which is consistent with the seasonal pattern of reported dengue infections in several of these regions such as southern Brazil, northern Australia, northern India, southern China, and parts of Central America. Temperate environments in the southern United States and arid regions across West Africa were estimated to have particularly limited windows of transmission (i.e. lower persistence suitability), despite comparatively high transmission suitability when averaged across the years. In other words, many regions can maintain brief seasons with high transmission suitability that permit short-lived epidemic activity but are unlikely to support year-round endemicity.

### Long-term trends in climate-based transmission suitability

To identify areas where transmission suitability has statistically significantly changed in response to climate change over the last 40 years (from now on referred to as *climate stress*), we quantified long-term trends in monthly time series of suitability per spatial pixel (see *Materials and Methods*; **Fig. 2)**. Climate stress towards both higher and lower transmission suitability was concentrated within the tropical and subtropical zones (**Fig. S3C**) with a steep decline in the presence and magnitude of stress at greater latitudes (**Fig. S5**). This spatial distribution coincided with regions that have historically exhibited high transmission suitability and encompassed much of South America, Africa, and Southeast Asia, where we estimated that 25.8%, 28.5% and 58.5% of land area have experienced climate stress over the past 40 years, respectively.

**Fig. 2.**
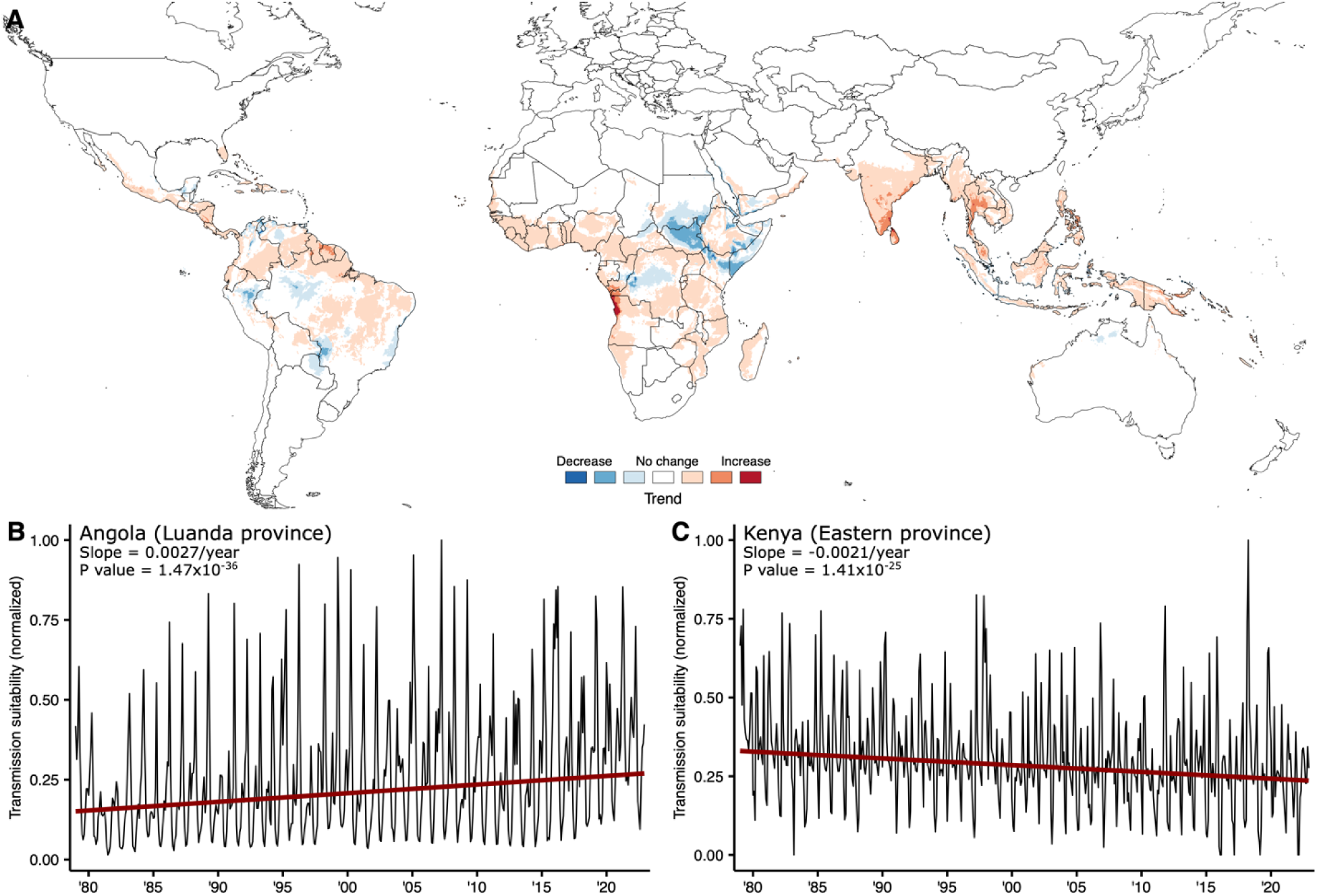
Global map of historical DENV climate stress. (**A**) Estimated yearly change in climate-based dengue transmission suitability (i.e. climate stress) per spatial pixel from 1979-2022 using the seasonal Mann-Kendall (MK) trend test and Sen’s slope on monthly Index P time series (dark blue = (−, −0.03], medium blue=[−0.03, −0.01), light blue=[−0.01, −0.005), white=[−0.005, 0.005), light orange=[0.005, 0.01), medium orange=[0.01, 0.03), dark orange=[0.03,)]. Spatial pixels with trends that do not meet FDR<0.05 are also colored white. (**B**, **C**) Examples of long-term trends in transmission suitability for spatial pixels in Luanda province, Angola and Eastern province, Kenya. Monthly Index P time series (black line) and trend estimates (red line) are shown. Results from the seasonal MK trend test including the p-value and Sen’s slope are provided. Y-axis is normalized by the maximum value.

By measuring historical changes in the land area with a high suitability for transmission, we examined how and where the geographical limits of transmission have been affected by climate stress (**Fig. 3**). Since climate stress was predominantly identified in areas with historically high transmission suitability, we estimated relatively small changes in the total land area at high risk of transmission. Globally, 11.5% of total land area was estimated to be under climate stress, with the high-risk land area increasing from 15.6% to 16.6% (Δ=1.1 percentage points (pp), 90% credible interval (CI)=0.7-1.6). (**Fig. 3B**). Similarly, the total high-risk land area marginally increased from 33.0% to 35.9% (Δ=2.9 pp, 90% CI=0.1-5.3) in Africa, and from 8.8% to 10.0% (Δ=1.2 pp, 90% CI=0.7-2.3) in Asia (**Fig. 3B**).

**Fig. 3.**
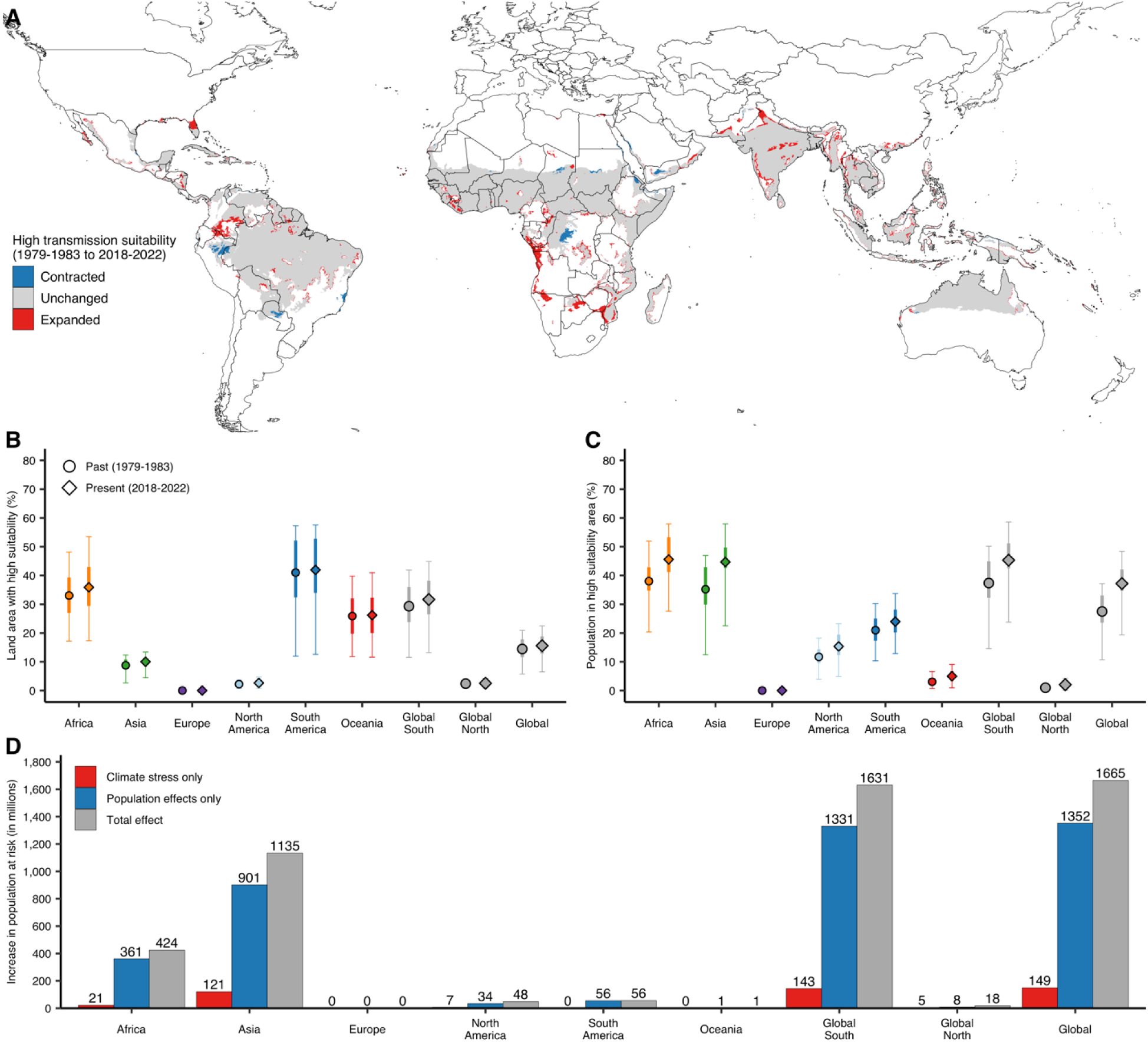
Estimated changes in land area and population with high climate-based DENV transmission suitability. (**A**) Changes in areas classified as having high transmission suitability (using a threshold of 1.0). (**B**, **C)** Estimated changes in land area (**B**) and population (**C**) with high transmission suitability from 1979- 1983 (circle) to 2018-2022 (diamond) for different regions. Mean (point), 50% (thick bar) and 90% (thin bar) credible intervals are shown. (**D**) Increase in the population (in millions) living in areas with high transmission suitability from 1979-1983 to 2018-2022 due to climate stress (red), demographic change (blue) and both climate stress and demographic change (grey). Spatial pixels with trends that do not meet FDR<0.05 were classified as having no change in transmission suitability.

Locally, however, there was substantial spatial heterogeneity **(Fig. S6-S7, Table S1)**. Significant climate stress was concentrated in the Global South where transmission suitability has been historically high, with relatively little change in the geographical limits of dengue transmission suitability in the Global North. For example, we estimated that the total high-risk land area increased by 5.1 pp (90% CI=0.1-8.1) from 37.0% to 42.1% across Central Africa and by 6.1 pp (90% CI=4.4-7.3) from 55.4% to 61.5% across Southeast Asia. Similarly, several countries with high numbers of reported DENV cases across Southeast Asia showed large changes in total high-risk land area, including Malaysia (Δ=8.4 pp), the Philippines (Δ=8.5 pp), Thailand (Δ=10.1 pp) and Myanmar (Δ=11.4 pp).

Significant negative trends in transmission suitability were identified in some parts of Eastern Africa and South America (**Fig. 2, Fig. S8**). This might reflect gradual changes in temperature and humidity beyond the optimal thermal ranges required to support *Ae. aegypti* transmission. Nonetheless, these areas remain highly suitable for DENV transmission, highlighted both by our estimates of transmission suitability (**Fig. 1**) and evidence of recent DENV epidemic activity (*27*). We did not identify significant climate stress in Europe, North America, Australia or north Asia over the last four decades. However, some isolated areas within these larger regions have shown signs of climate stress towards higher suitability. These included coastal areas of Spain and Italy in Europe, Queensland in Australia, and Florida in the United States of America (**Fig. S9-S11**, **Table S1**).

### Global change in human populations in high-risk areas

We further examined how human demographic changes (e.g. population growth and migration) coincided with the estimated distribution of climate stress towards higher transmission suitability (**Fig. 3, Table S2**). We estimated that the proportion of the global population living in high-risk areas increased from 27.5% to 37.2% (Δ=9.7 pp, 90% CI=8.5-12.3; **Fig. 3C**), resulting in 1.67 billion more people living in areas with high-risk climate conditions for dengue transmission (**Fig. 3D**).

The majority of this trend was driven by changes in Africa where the proportion of the population in high-risk settings increased from 38.0% to 45.6% (Δ=7.6 pp, 90% CI=5.1-10.6) and in Asia where the proportion increased from 35.2% to 44.7% (Δ=9.5 pp, 90% CI=6.8- 14.3) (**Fig. 3C**). In absolute terms, these changes represent 424 million and 1.14 billion more people living in high-risk areas in Africa and Asia, respectively. In Asia, four countries (India, Indonesia, China and Bangladesh) contributed over 80% of the estimated increase (**Table S2**). In Africa, the increase was more uniformly distributed with Western Africa, especially Nigeria, responsible for the greatest share of this increase (**Table S2**).

The estimated change in the share of the population living in high-risk areas was comparatively smaller across the Americas with an increase from 11.7% to 15.3% (Δ=3.6 pp, 90% CI=1.1-5.6) across North America (roughly 48 million more people), and from 21.0% to 24.0% (Δ=3.0 pp, 90% CI=2.3-3.5) across South America (roughly 56 million more people) (**Fig. 3C-D**). In South America, we estimated that Bolivia, Brazil, Columbia, Ecuador and Venezuela had among the largest increases in the share of the population living in high-risk areas (**Table S2**). Substantial changes were also estimated for countries across Central America and the Caribbean, including Haiti, El Salvador, and the Dominican Republic, where the share of the population in highly favorable climate conditions increased by over 10 percentage points. (**Fig. 3C, Table S2**).

To estimate the independent contribution of climate and demographic changes to these increases in the population living in high-risk areas, we considered hypothetical scenarios in which climate and demographic effects were each held constant over the four decades (**Fig. 3D**). Globally, we estimated that population growth contributed to the vast majority of the estimated increase in the population living in high-risk areas. Demographic and climate changes independently contributed 1.35 billion and 149 million more people in high-risk areas, respectively. This observation held across continents (except Europe where there were no changes in the number of people at high risk), with demographic changes responsible for at least 5-fold more people living in high-risk areas than that estimated for the effects of climate stress alone. Hence, while the total population living in areas suitable for DENV transmission has increase, the contribution of climate change to this phenomenon pales in comparison to that of population growth and migration in areas classically associated with DENV transmission.

When we accounted for the combined effects of climate and demographic change, we observed that the summation of the independent effect of each factor was less than their combined effects (1.50 billion vs. 1.67 billion; **Fig. 3D**). This suggests that population growth frequently occurred in locations which have only recently exceeded the high suitability threshold. Furthermore, the greater increase in the proportion of the global population living in high-risk areas (from 27.5% to 37.2%; **Fig. 3C**) compared to the increase in the total high-risk land area (from 15.6% to 16.6%; **Fig. 3D),** indicates that population growth has also disproportionately occurred in areas with historically high transmission suitability.

### Shifts in transmission suitability in densely populated areas

Since *Ae. aegypti* prospers within densely-populated urban environments, we examined and summarized how transmission suitability changed in areas with different human population density over the past 40 years (**Fig. 4**). Globally, the proportion of high and very high density areas with high-risk conditions for DENV transmission increased from 26.9% to 36.5% (Δ=9.6 pp, 90% CI=8.0-13.4) and from 20.2% to 35.4% (Δ=15.2 pp, 90% CI=10.7-17.4), respectively.

**Fig. 4.**
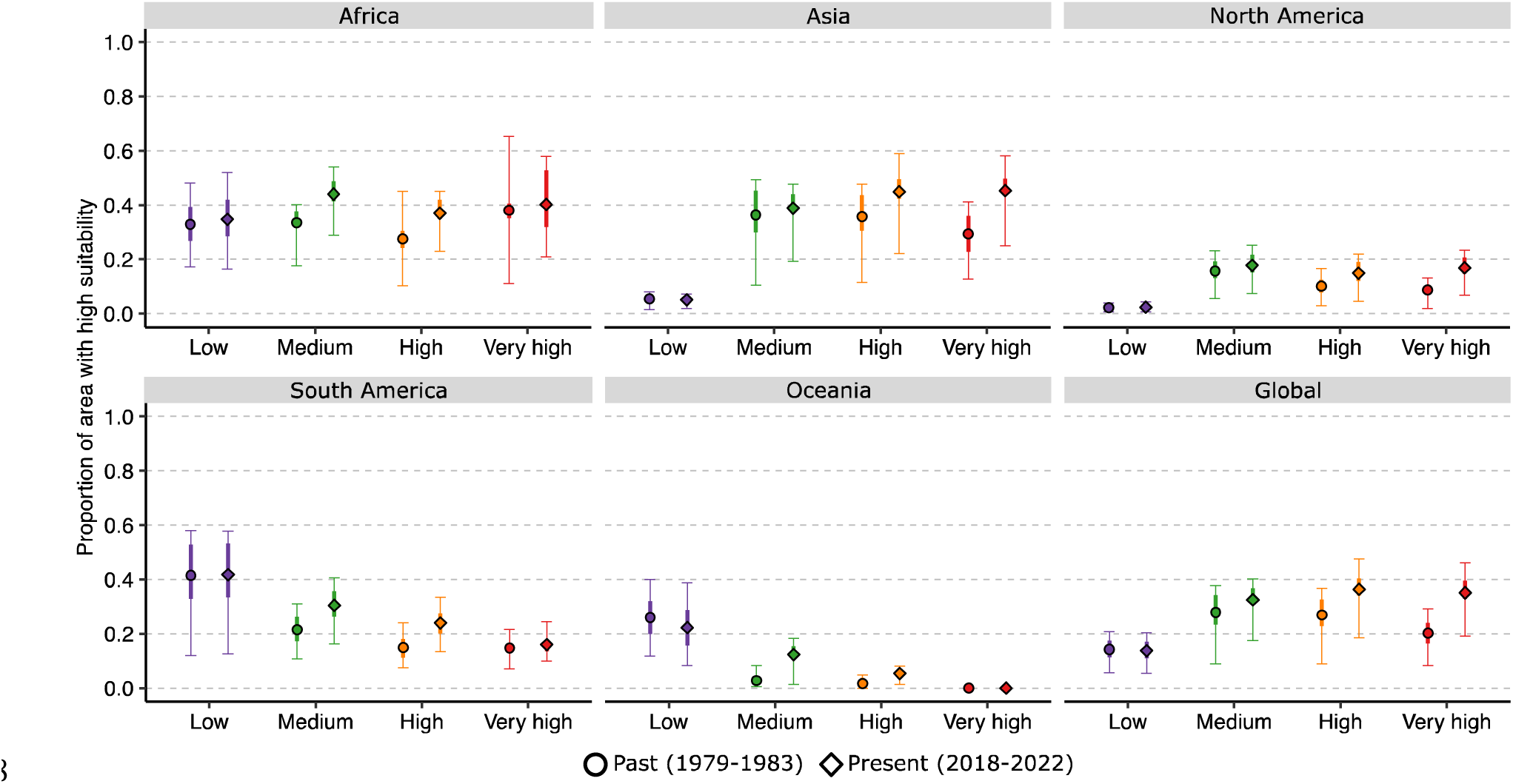
Shifts in the land area with high climate-based DENV transmission suitability stratified by population density. The proportion of land area with climate suitable for DENV transmission (using a threshold of 1.0) stratified by population density in 1979-1983 (circle) and 2018-2022 (diamond). The population density of spatial pixels was divided into low density (<100 people/km^2^), medium density (101-300 people/km^2^), high density (301-1500 people/km^2^) and very high density (>1501 people/km^2^). Mean (point), 50% (thick bar) and 90% (thin bar) credible intervals are shown. Europe is not included because there were no estimated changes in the land area with high transmission suitability. Spatial pixels with trends that do not meet FDR<0.05 were classified as having no change in transmission suitability.

In contrast, the proportion of low and medium density areas with high-risk conditions were roughly unchanged over the last 40 years. Again, these results suggest that high density population centers have increasingly concentrated in areas where transmission suitability for DENV is historically favorable. This global trend, however, masked regional heterogeneity in the magnitude of these changes. For example, we estimated relevant increases in the proportion of both high and very high density areas with high-risk conditions for DENV transmission across Asia and North America (**Fig. 4**). Conversely, we estimated little change in the proportion of very high density areas with favorable climate conditions across Africa, South America and Oceania, where we instead observed consistent increases in the proportion of medium and high density areas with high transmission suitability.

## Discussion

In this study we used a DENV suitability measure that incorporates the effects of temperature and humidity on mosquito-viral traits to evaluate long-term changes in transmission suitability at a global scale over the last 40 years. We contrasted these long-term trends in transmission suitability with changes in the size and distribution of the global population. Our findings indicate that historical climate stress has mostly occurred in the equatorial tropical and subtropical zones spanning much of Sub-Saharan Africa, Southeast Asia, the Western Pacific and northern Latin America. Although the effects of climate stress varied considerably among regions, the last four decades were marked by a substantial increase in the size of human populations living in areas with climate conditions highly favorable to DENV transmission.

Climate stress was predominantly concentrated in areas that have historically exhibited both high transmission suitability and epidemic activity, resulting in only marginal changes to the geographical limits of suitability. There was, however, regional heterogeneity in the distribution of climate stress, with the Global South accounting for much of the expansion in land area with high transmission suitability. Several of the areas with marked increases in suitability due to climate stress have recently reported increased local transmission (e.g. southern United States (*28*) and Central Africa (*27*)) or larger and longer seasonal outbreaks (e.g. Central America (*29*)). In contrast to many existing future predictions of increased dengue risk in continental Europe (*30*), we found that transmission suitability has remained relatively low across Europe over the last 40 years, with increases in transmission risk confined to coastal areas in Spain, France, Italy and Turkey. Some of these areas, specifically Southern Spain, France and Northern Italy have seen a rise in short-term autochthonous transmission of dengue, chikungunya and West Nile virus in the past 5 years (*31*, *32*).

Interestingly, some regions, most notably Eastern Africa, are estimated to be in the process of long-term decreases in transmission suitability. This suggests that the increasingly dry and warm conditions in regions within and around Eastern Africa may be negatively affecting mosquito-viral traits, resulting in decreased overall DENV transmission suitability. These results point to a potentially ongoing, long-term shift in the geographical distribution of dengue risk with the emergence of DENV in new areas and its disappearance from classically suitable areas in the future. This observation is consistent with future projections of dengue risk (*6*) and may follow the expected climate-driven decline in the transmission of malaria by the *Anopheles gambiae* mosquitoes, which are adapted to cooler conditions, in Africa (*33*, *34*). Many of these areas with decreasing suitability trends, however, remain climatically-suitable for dengue transmission, emphasizing that climate-driven constraints on dengue transmission may not materialize for many years and are still subject to future climate change (*35*).

When we examined the interaction between transmission suitability and recent global demographic changes we found that population growth has preferentially occurred in areas with historically high suitability, resulting in substantial increases in the population living in high-risk areas. Assuming favorable conditions for the establishment of vector populations (e.g. suitable reproduction habitat), close to 50% of the populations in Africa and Asia are currently living in areas with a high risk of infection - a 25% increase relative to four decades earlier. Therefore, despite relatively small changes in the geographic limits of transmission suitability, parallel demographic changes have resulted in millions of additional people living in high-risk areas. This rapid growth in the available host population in climate-suitable areas emphasizes the increasing public health burden that the Global South will face as DENV continues to spread to new areas.

The increase in the proportion of high and very high human density areas with favorable climate conditions has implications for how we understand the recent increase in global dengue incidence. In a recent study on future dengue risk in Southeast Asia, Colón-González et al. (*7*) found that high density areas, relative to both low and very high density areas, confer the highest risk of infection. Taken together with their findings, our results point to the emergence of high density population centers as a potential contributor to the currently high risk of DENV transmission in many parts of the world. Additionally, many epidemiological studies from dengue-endemic areas in Southeast Asia have identified an association between poor urban planning (i.e. limited water infrastructure and waste management) and increased dengue transmission (*36*, *37*). Hence, there is a need to more closely examine the link between urban development in high density areas and climate-based dengue risk. These links are likely to become increasingly important as the Global South, particularly Africa and South America, continues to undergo rapid urbanization and industrial development while facing higher dengue transmission suitability.

There are several limitations as well as potential future opportunities of this study. Our model considers the empirical effects of temperature and humidity on transmission suitability, but other factors including precipitation (*8*, *10*), altitude (*38*), urbanization (*37*), and vegetation (*8*) could either constrain or favor local transmission. Furthermore, our suitability index does not provide any assurance that the mosquito vector, human host, or the virus are present and instead assumes that all three are present. Although we have previously demonstrated that the index robustly reproduces spatiotemporal patterns of dengue incidence (*26*), high climate-based transmission suitability (i.e. Index P) should not be interpreted as a guarantee of dengue epidemic activity, but rather as the fulfillment of one of several criteria necessary for DENV transmission. For areas without reported dengue cases, our transmission suitability maps can help identify hotspots where there is climate-based potential for introduction or where incidence might be under-reported due to insufficient surveillance. For example, we estimated high transmission suitability in Central Africa where *Ae. aegypti* is predicted to be widely disseminated (*8*) but reported cases remain low (*27*). We also identify areas in Europe and USA with non-significant but probable suitability changes in the last 40 years that have recently witnessed viral introductions albeit with short-term transmission success. Conversely, large differences in estimated transmission suitability and reported cases were observed in arid environments such as central Australia where suitable mosquito breeding habitats may be scarce (e.g. due to low precipitation). Similarly, differences between dengue reporting and transmission suitability may arise in sparsely populated areas such as the Amazon in South America where barriers to human and viral mobility, as well as necessary human density for sustained transmission, likely limit the occurrence of chains of transmission.

The mathematical expressions for mosquito-viral traits used in our suitability index are based on experimental data from *Ae. aegypti* mosquitoes and do not account for the potential contribution of *Ae. albopictus* mosquitoes to DENV transmission. This would be relevant e.g. for southern Europe, where *Ae. albopictus* is now established but *Ae. aegypti* is not. Previous research has shown that adult *Ae. albopictus* have higher survival rates than *Ae. aegypti* and are better adapted to persist at higher latitudes (*9*, *15*). Using these experimental results, Brady et al. (*15*) developed maps defining the current thermal limits of persistence for these two vectors. It could be informative to contrast the historical geographical distribution of transmission suitability for these vectors while including the influence of both temperature and humidity. However, currently available empirical data on mosquito-viral traits as a function of climate variables do not provide sufficient information to fully parameterize and robustly validate Index P specific for *Ae. albopictus*.

One of the major challenges of using our results to inform dengue control efforts will be to understand how favorable climate-based transmission suitability may drive future dengue transmission in specific locations and contexts. While we have previously characterized the relationship between Index P and dengue incidence across a variety of environmental conditions (*26*), we encourage researchers to interpret our estimates for a region of interest within the context of local demographic, economic and ecological conditions. Furthermore, our estimates of transmission suitability reflect the past and should not be interpreted as predictions for what will happen decades into the future. Given recent acceleration in global warming, projections of future dengue risk based on climate change forecasts may significantly differ from the distribution and magnitude of climate stress reported in this study.

In this study, we aimed to better understand how the current spatiotemporal epidemiology of dengue has been shaped by historical climate and demographic changes. The reported outputs stand as a complement to a number of other studies focused on mapping the distribution of DENV vectors (*8*) and forecasting future transmission based on climate change projections (*6*, *7*). Overall we demonstrate that climate stress can have both detrimental and beneficial effects on dengue transmission suitability, and that while historical climate stress has contributed to an overall increase in global transmission suitability, parallel demographic changes have been instrumental in expanding the population living in areas with a high risk of dengue transmission.

## Materials and Methods

### Index P theory

Briefly, the basic reproduction number R_0_ of a mosquito-borne virus can be summarized as the product of two factors: the average number of adult female mosquitoes per host (M) and the transmission potential of each adult female mosquito (P) in an immunologically naive host population. Both M and P oscillate in time in response to environmental and ecological factors. Due to an absence of high-resolution data on mosquito density, M is often unknown or difficult to parametrize. The transmission potential P depends on host, viral and mosquito traits (e.g. incubation periods and lifespans), several of which have been shown to depend on meteorological variables including temperature (*9*, *14*) and relative humidity (*16*, *17*). Since it is often challenging to mechanistically model M at a fine spatial and temporal scale, many theoretical epidemiological studies have estimated transmission potential using proxy measures based on known functions of the parameters that compose the basic reproduction number (*15*, *18*, *19*, *21*, *39*). These proxy measures are often referred to as “suitability indices’’, and while incomplete in terms of the inclusion of parameters known to be important for viral transmission, can offer reliable estimates of the real transmission potential of the mosquito-borne virus. In recent years, significant work has been done to validate their capacity to explain the spatio-temporal dynamics of mosquito-borne viruses (*21*, *26*).

In this study, we used Index P (derived from R_0_=MP) as a suitability index for climate-based DENV transmission potential by *Ae. aegypti* mosquitoes (*19*, *26*). Index P uses mathematical expressions for relationships between DENV-*Ae. aegypti* traits and meteorological variables (i.e. temperature and relative humidity) obtained from empirical studies, Thus, it includes climate-dependent functions of relevant entomological parameters such as the extrinsic incubation period, mosquito lifespan and mosquito biting rate. Longitudinal temperature and relative humidity time series are then used with these functions within a Bayesian sampling framework to derive estimates for P over time. Full methodological details and technical validation of Index P can be found in Obolski et al. (*19*) and Nakase et al (*26*). Index P has been successfully applied in several studies to characterize the transmission dynamics of dengue (*19*, *25*, *26*), chikungunya (*40*) and West Nile virus (*41*, *42*). In this current study, we use the same approach as described in Nakase et al (*26*) with the same epidemiological priors and climate-dependent functions for DENV transmitted by *Ae. aegypti* mosquitoes as applied and validated in that study.

### Spatiotemporal maps of climate-based transmission suitability

Spatiotemporal maps of climate-based transmission suitability for DENV spread by *Ae.* aegypti mosquitoes (i.e. Index P) were obtained from Nakase et al (*26*). We used the available dataset “Global climate-driven transmission suitability maps for dengue virus transmitted by Aedes aegypti mosquitoes from 1979 to 2022” (updated post-publication of the research article) which includes Index P time series for spatial pixels at a time resolution of 1 month (1979–2022) and spatial resolution of 0.25° x 0.25° for 186 countries and territories.

### Summarization of spatiotemporal maps of suitability

To minimize the effect of outlier years and align the Index P data with the available population data, we used the average Index P from 1979 to 1983 and from 2018 to 2022 as estimates of past and present climate-based DENV transmission suitability. We calculated climate suitability for DENV persistence by averaging the number of months Index P was above a threshold of 1.0 per year from 1979 to 1983 and from 2018 to 2022, respectively. An Index P threshold of 1.0 was selected because it corresponds to the basic reproduction number of 1.0 in a population where the average number of adult female mosquitoes per host is 1.0. Uncertainty estimates including the 50% and 90% credible intervals for these values were calculated using summary statistics from 1000 simulations of the spatiotemporal maps of Index P provided with the dataset from Nakase et al (*26*).

### Analysis of long-term suitability trends

Long-term transmission suitability trends were estimated using the seasonal Mann-Kendall (MK) test (*43*) on the monthly Index P time series from 1979 to 2022. The MK test for trend detection is a non-parametric method based on rank that tests for the existence of a monotonic continuous trend in serially independent data. We use the seasonal extension of the MK test on the monthly Index P time series (528 data points for each spatial pixel) to account for the observed seasonality in the transmission potential during a calendar year. For each spatial pixel, we applied the seasonal MK test with statistical significance defined by a false discovery rate (FDR) of 5% across all pixels. We then used Sen’s slope (*44*) to estimate the linear rate of change for those spatial pixels with statistically significant trends. Slope estimates along with associated p-values are provided in **Fig. S12**.

### Population data

Global gridded population counts and population density for 1980 and 2020 were obtained from the Socioeconomic Data and Applications Center (SEDAC) database (*45*, *46*) and the WorldPop database (*47*) respectively at a resolution of 30 arc (∼1km at the equator). The global gridded population data was aggregated to match the spatial resolution of the Index P maps using the *aggregate* function in the *raster* R package (*48*).

### Estimation of climate and demographic effects on population at risk

The independent effect of climate change on the population living in areas with high-risk suitability for DENV transmission was estimated by fixing the global population at the year 1980 and allowing climate-based DENV transmission suitability to change from 1980 to 2020. Similarly, the independent effect of demographic changes was estimated by fixing the climate-based DENV transmission suitability at the year 1980 and allowing the global population to change from 1980 to 2020.

## Data Availability

All data produced in the present work are contained in the manuscript.

## Author contributions

Conceptualization: TN, JL

Methodology: TN, UO, JL

Investigation: TN

Visualization: TN

Supervision: JL

Writing—original draft: TN, JL

Writing—review & editing: TN, MG, UO, JL

## Data and materials availability

All data needed to evaluate the conclusions in the paper are present in the paper and/or the Supplementary Materials. The raw global spatiotemporal maps of climate-based DENV transmission suitability from 1979-2022, which are the basis of these analyses, have previously been made publicly available on *figshare* (https://doi.org/10.6084/m9.figshare.21502614.v5).

## Supplementary Materials

**Supplementary Text File 1** includes Supplementary Figures S1-12.

**Supplementary Text File 2** includes Supplementary Tables S1-2.

