## Supplementary Text File 1 for "A retrospective analysis of climate-based dengue virus transmission suitability and demographic changes over the last four decades"

Taishi Nakase* *et al.*

**This PDF file includes:**

Figs. S1 to S12

**Other Supplementary Materials for this manuscript include the following:**
Tables S1 to S2

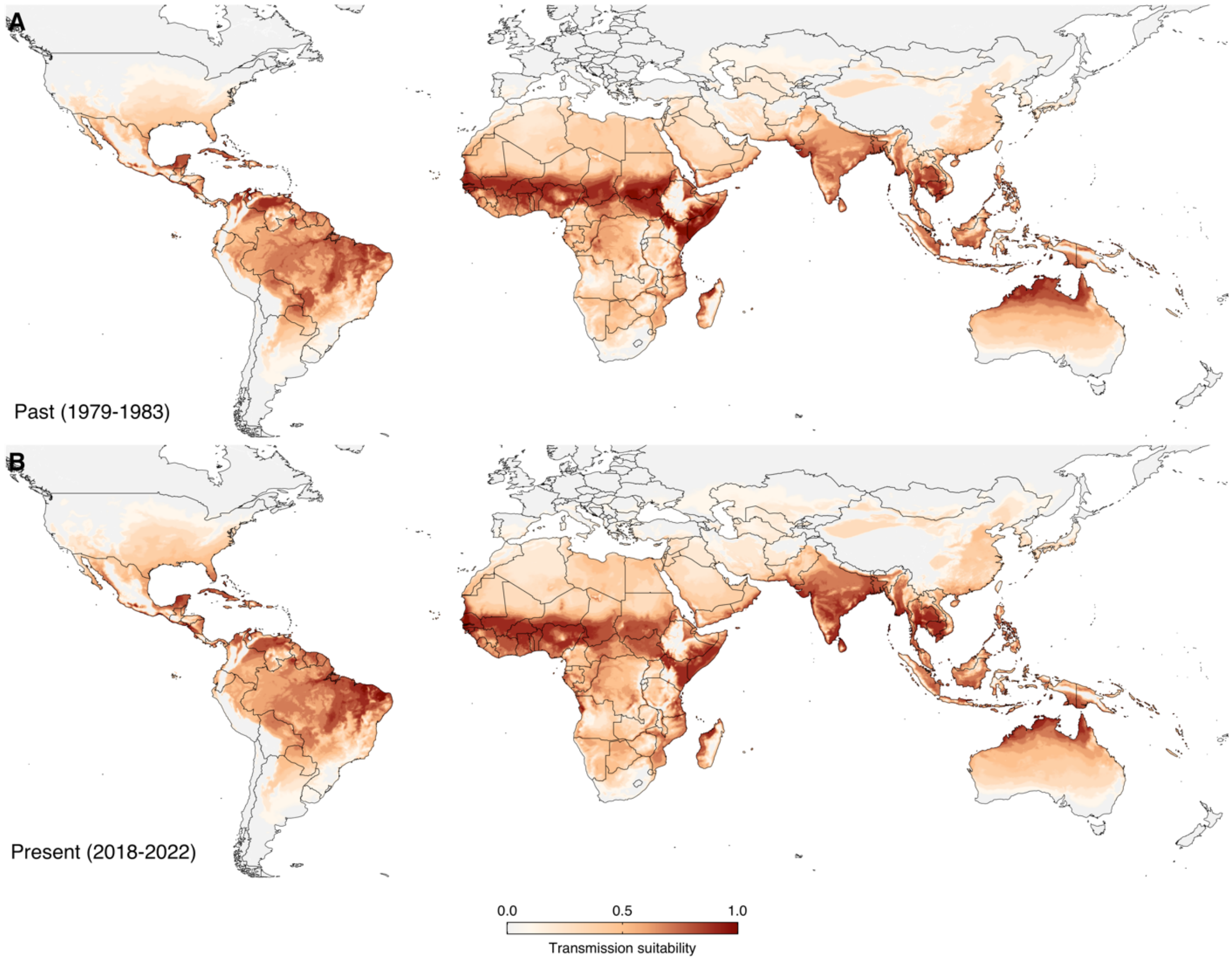


**Fig. S1. Global map of past and present DENV transmission suitability.** (**A**, **B**) For visualization purposes, average transmission suitability for 1979-1983 and 2018-2022 are presented on a logarithmic scale (normalized by maximum value). Transmission suitability is calculated by averaging the transmission potential over the respective time intervals.

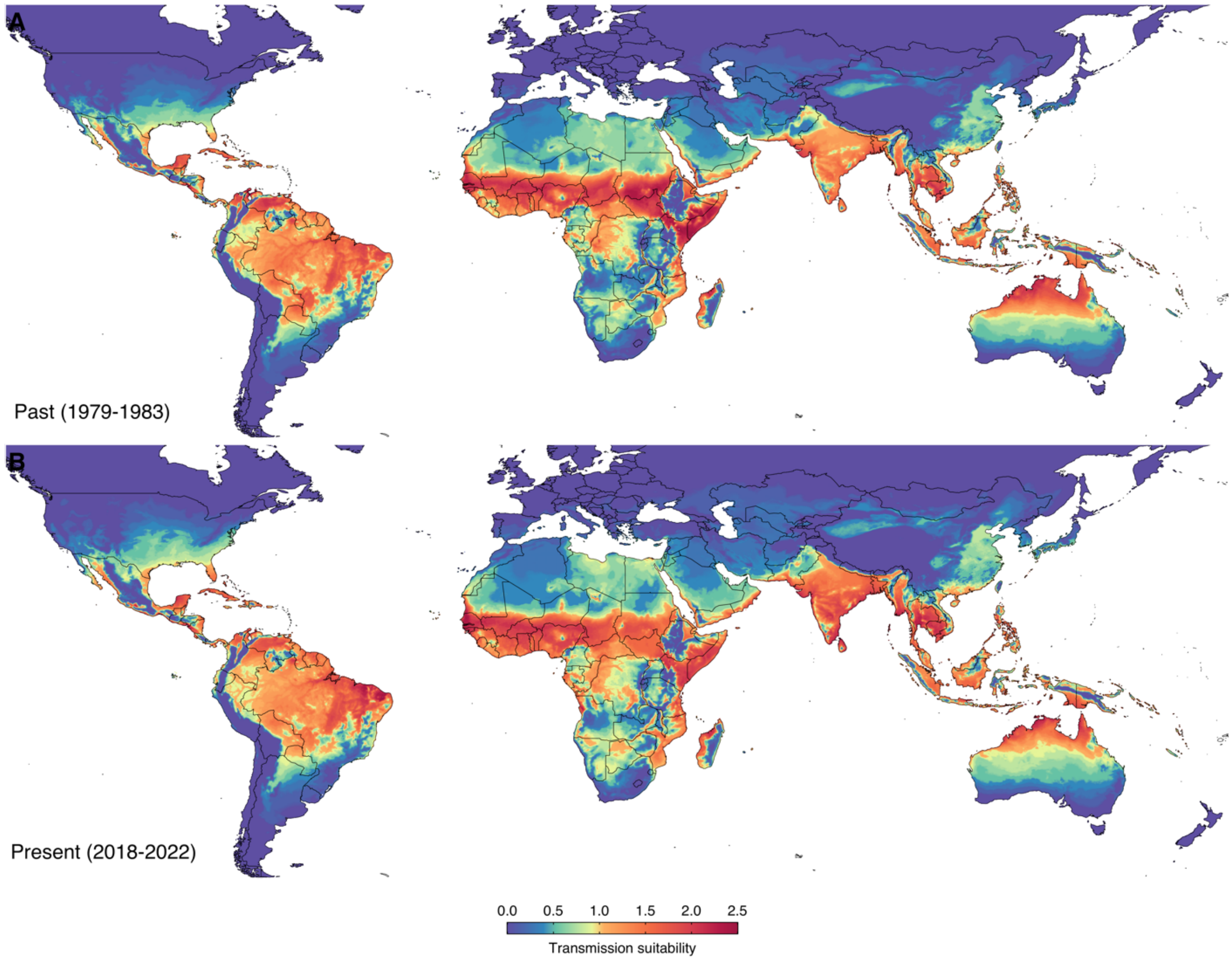


**Fig. S2. Global map of past and present DENV transmission suitability on absolute scale.** (**A**, **B**) Average transmission suitability for 1979-1983 and 2018-2022, respectively. Transmission suitability is calculated by averaging the transmission potential over the respective time intervals.

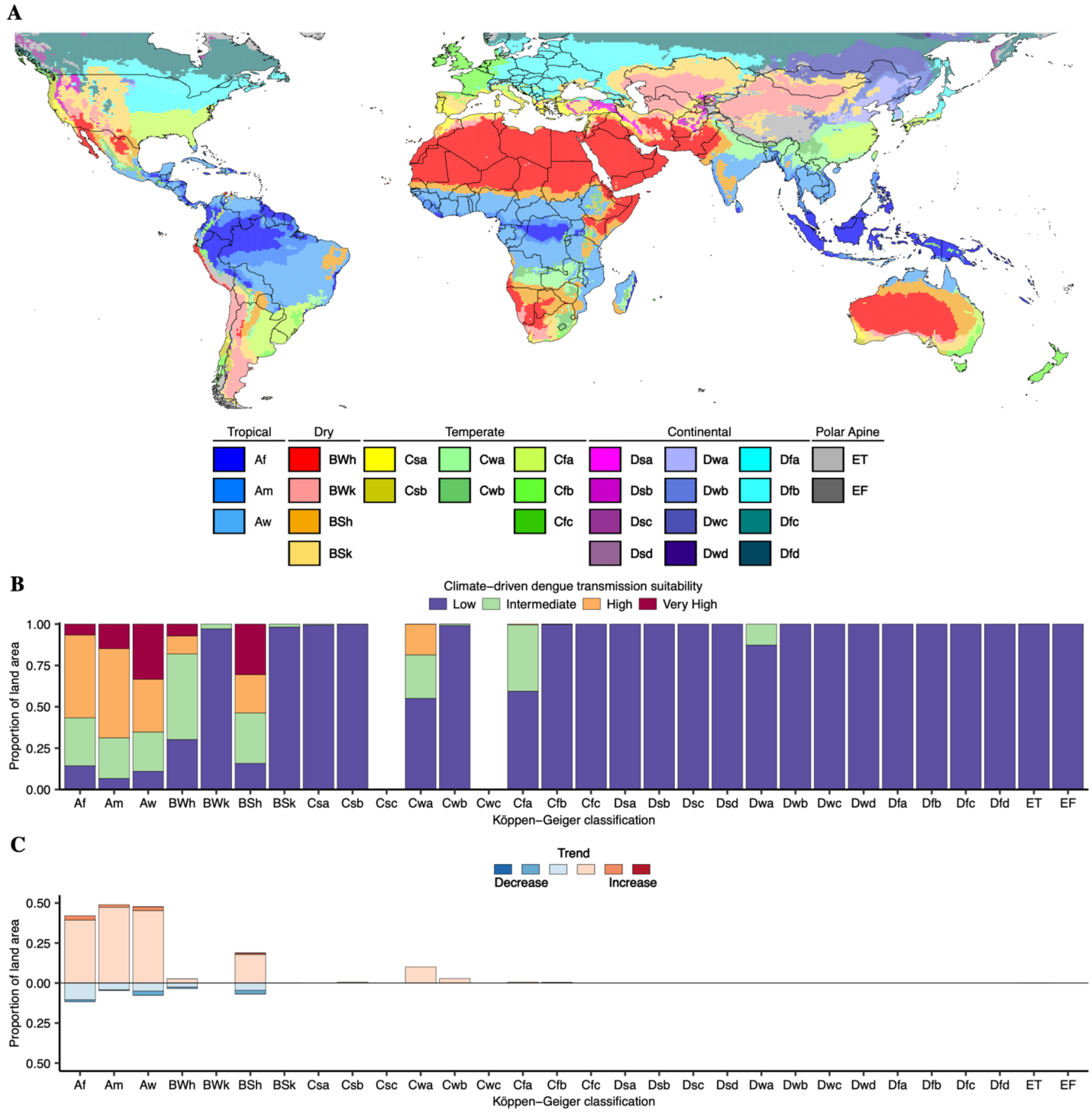


**Fig. S3. Historical trends in climate-based dengue transmission suitability stratified by the current Köppen-Geiger climate classification**. (**A**) Global map of the current Köppen-Geiger climate classification. (**B**) Average climate-driven DENV transmission suitability in 1979-1983 (low Index P = (0.0, 0.5); intermediate Index P = (0.5, 1.0); high Index P = (1.0, 1.5); very high Index P = (1.5, ∞)) stratified by the Köppen-Geiger climate classification. (**C**) Historical DENV climate stress from 1979-2022 stratified by the Köppen-Geiger climate classification. Same color palette as that used in Fig. 2A with white pixels not shown.

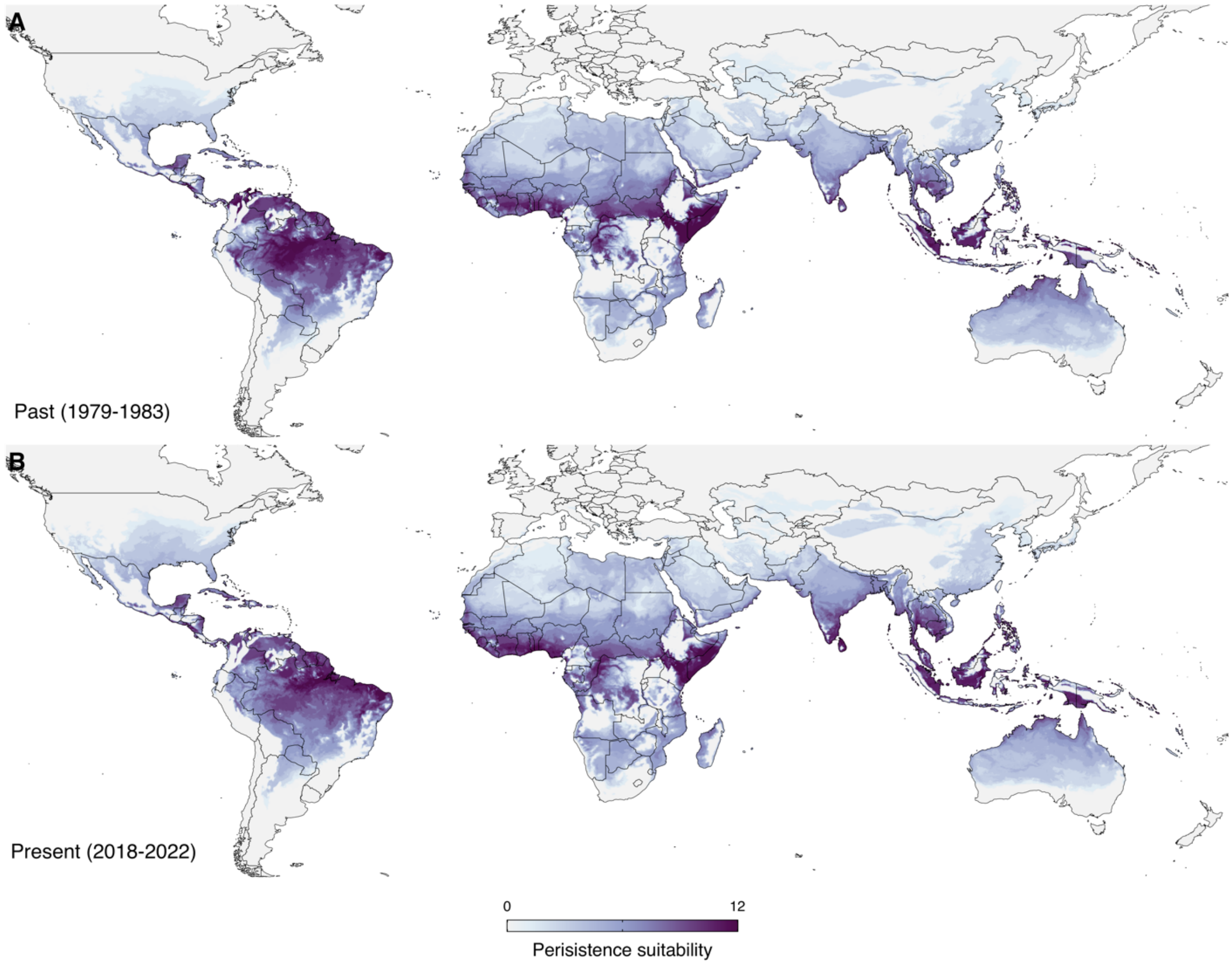


**Fig. S4. Global map of past and present DENV persistence suitability on absolute scale.** (**A**, **B**) Average persistence suitability (months) for 1979-1983 and for 2018-2022, respectively. Persistence suitability is measured as the number of months the transmission potential exceeds a threshold of 1.0 over a year, averaged over the respective time periods.

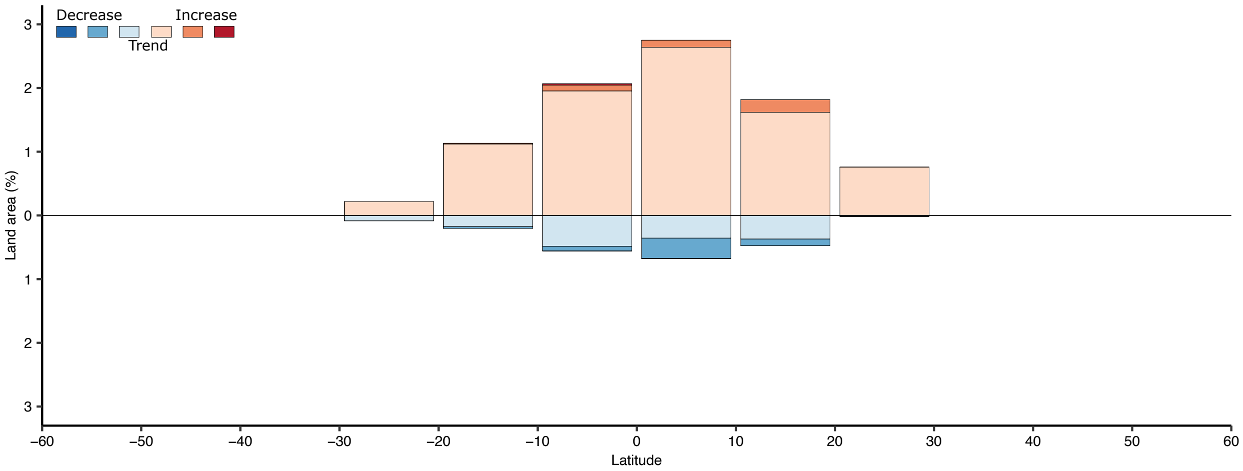


**Fig. S5. Historical DENV climate stress as a function of latitude**. Same color palette as that used in Fig. 2A with white pixels not shown.

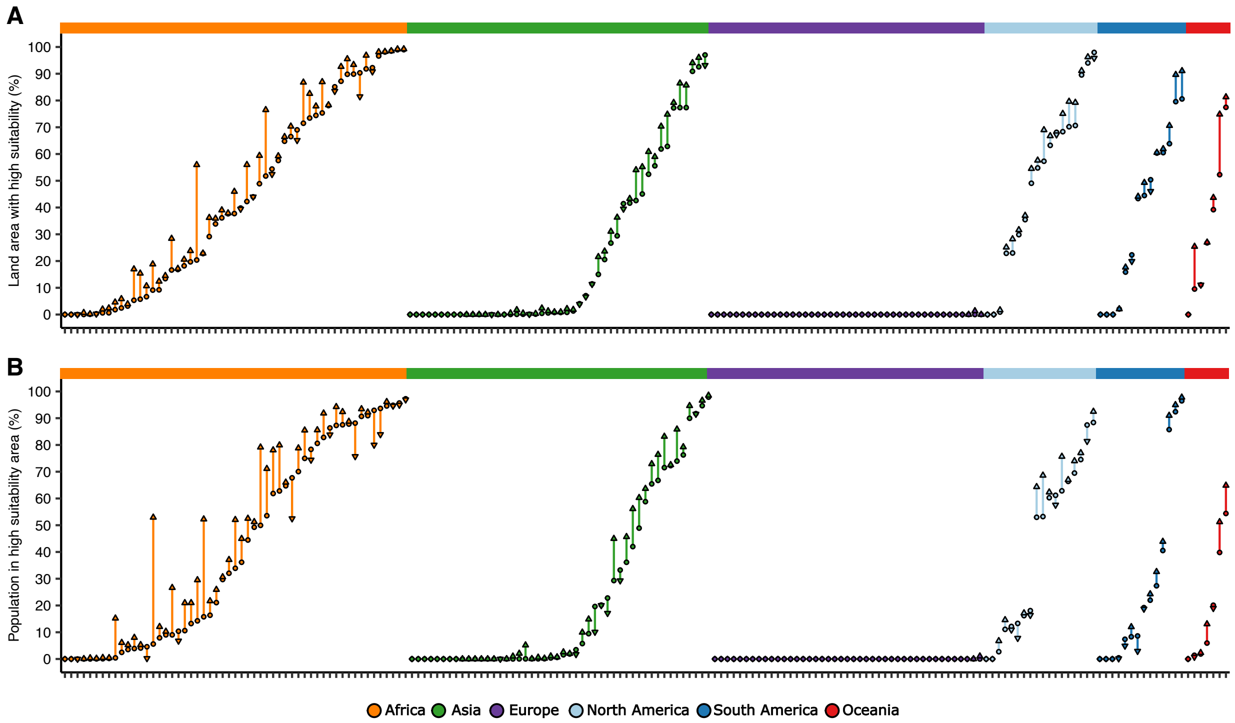


**Fig. S6. Estimated changes in land area and population with high climate-based DENV transmission suitability.** Estimated changes in land area (**A**) and population (**B**) with high DENV transmission suitability (using a threshold of 1.0) from 1979-1983 (circle) to 2018-2022 (downward triangle=decrease; diamond=no change; upward triangle=increase) for each country or territory. Spatial pixels with trends that do not meet FDR < 0.05 were classified as having no change in transmission suitability.

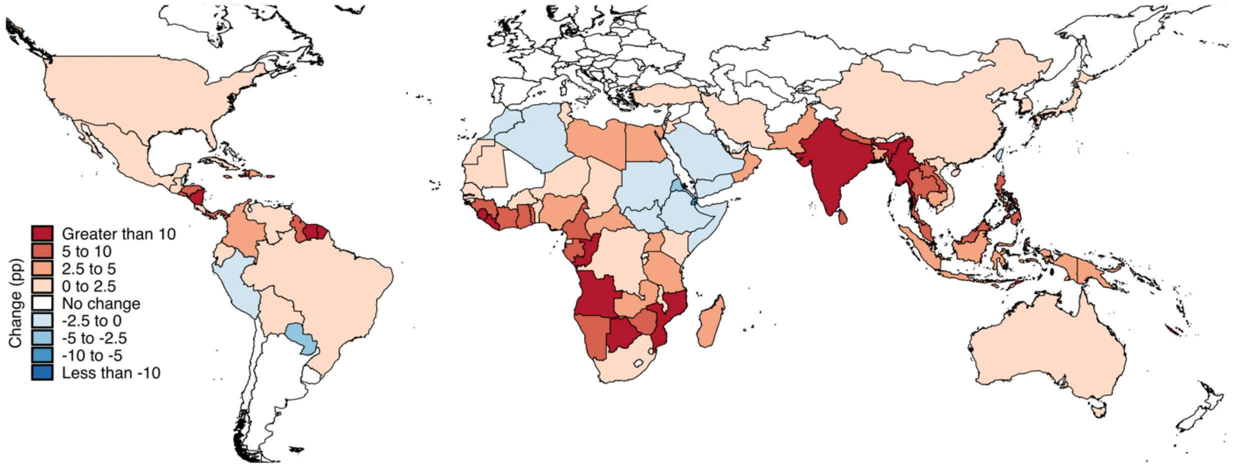


**Fig. S7. Change in the proportion of land area with high DENV transmission suitability from 1979-1983 to 2018-2022 per country/territory.** Spatial pixels with trends that do not meet FDR < 0.05 were classified as having no change in transmission suitability.

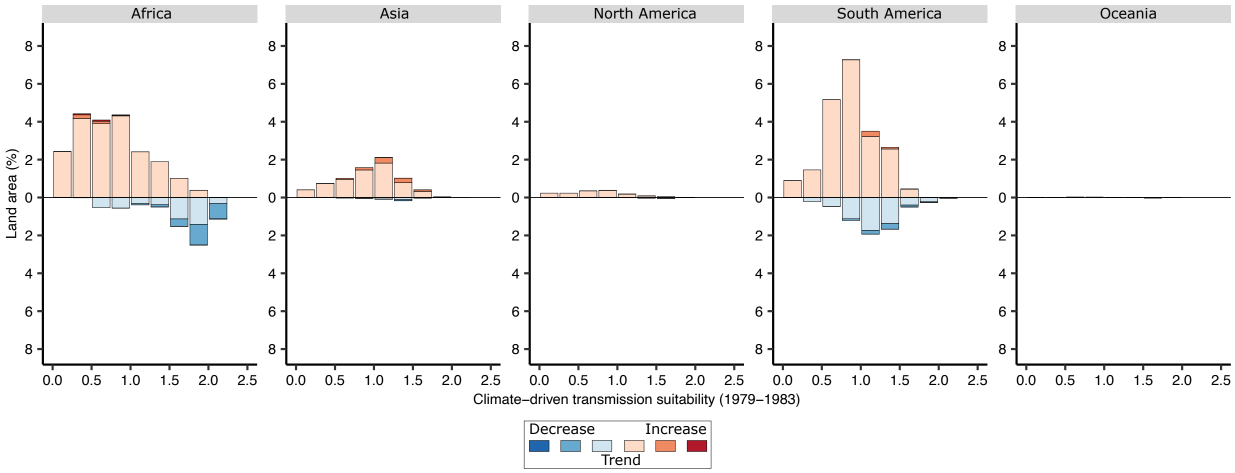


**Fig. S8. Historical climate stress as a function of past transmission suitability.** The y-axis represents the proportion of total land area across each continent under different levels of climate stress as a function of past transmission suitability in 1979-1983. Europe is not included because there were no pixels under significant climate stress. Same color palette as that used in Fig. 2A with white pixels not shown.

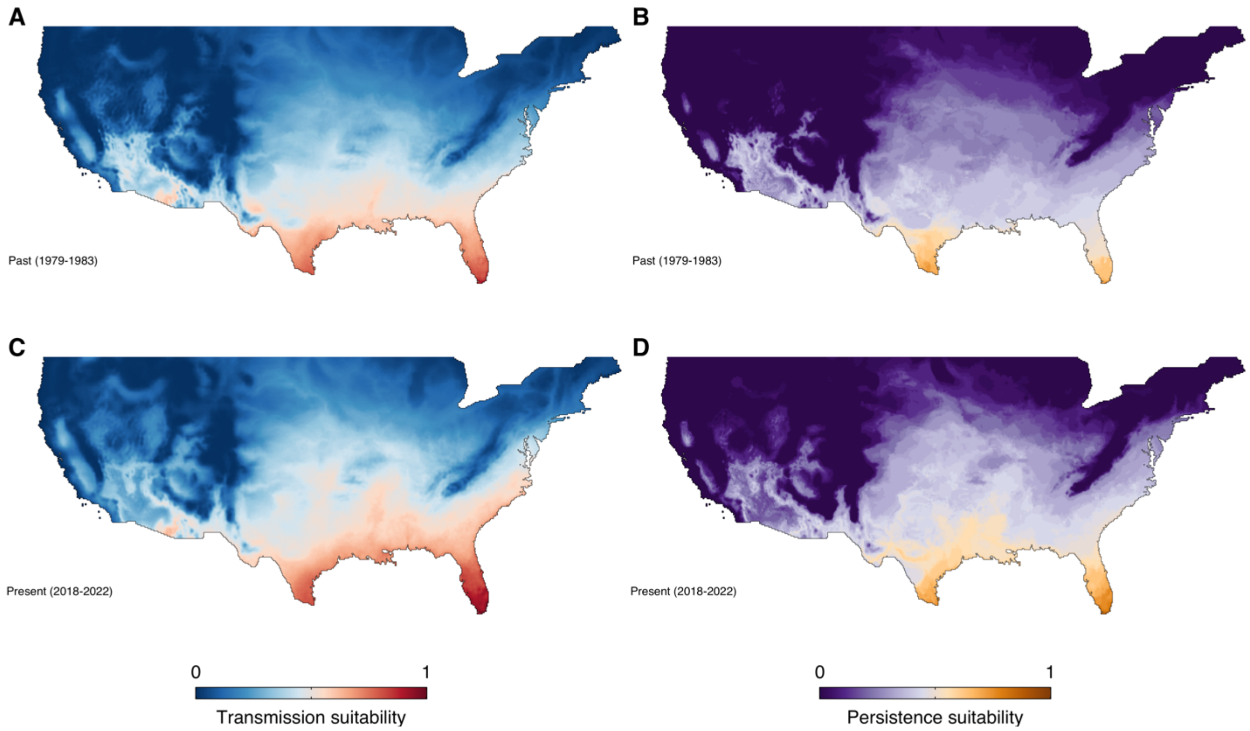


**Fig. S9. Map of past and present DENV transmission and persistence suitability in the United States.** (**A**, **C**). Average transmission suitability for 1979-1983 and 2018-2022 on a normalized scale, respectively. (**B**, **D**) Average persistence suitability for 1979-1983 and 2018-2022 on a normalized scale, respectively.

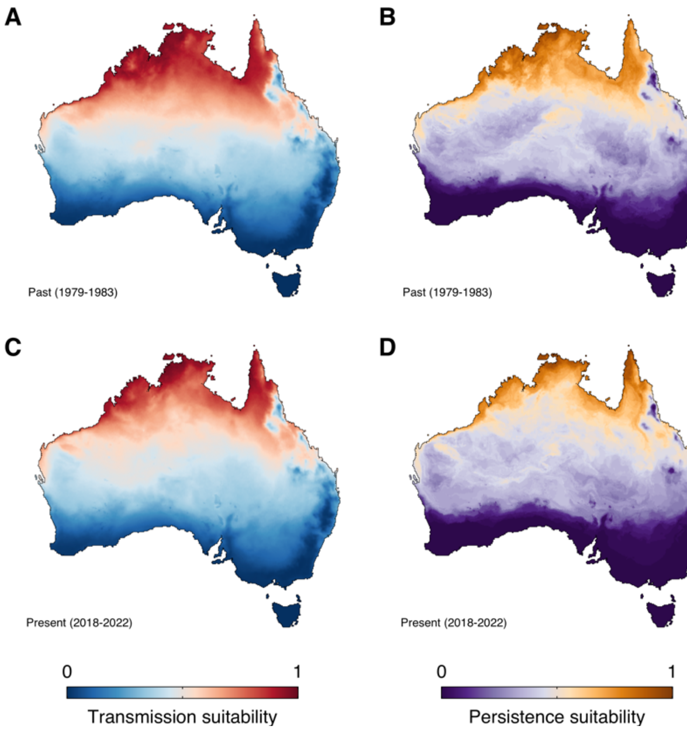


**Fig. S10. Map of past and present DENV transmission and persistence suitability in Australia.** (**A**, **C**). Average transmission suitability for 1979-1983 and 2018-2022 on a normalized scale, respectively. (**B**, **D**) Average persistence suitability for 1979-1983 and 2018-2022 on a normalized scale, respectively.

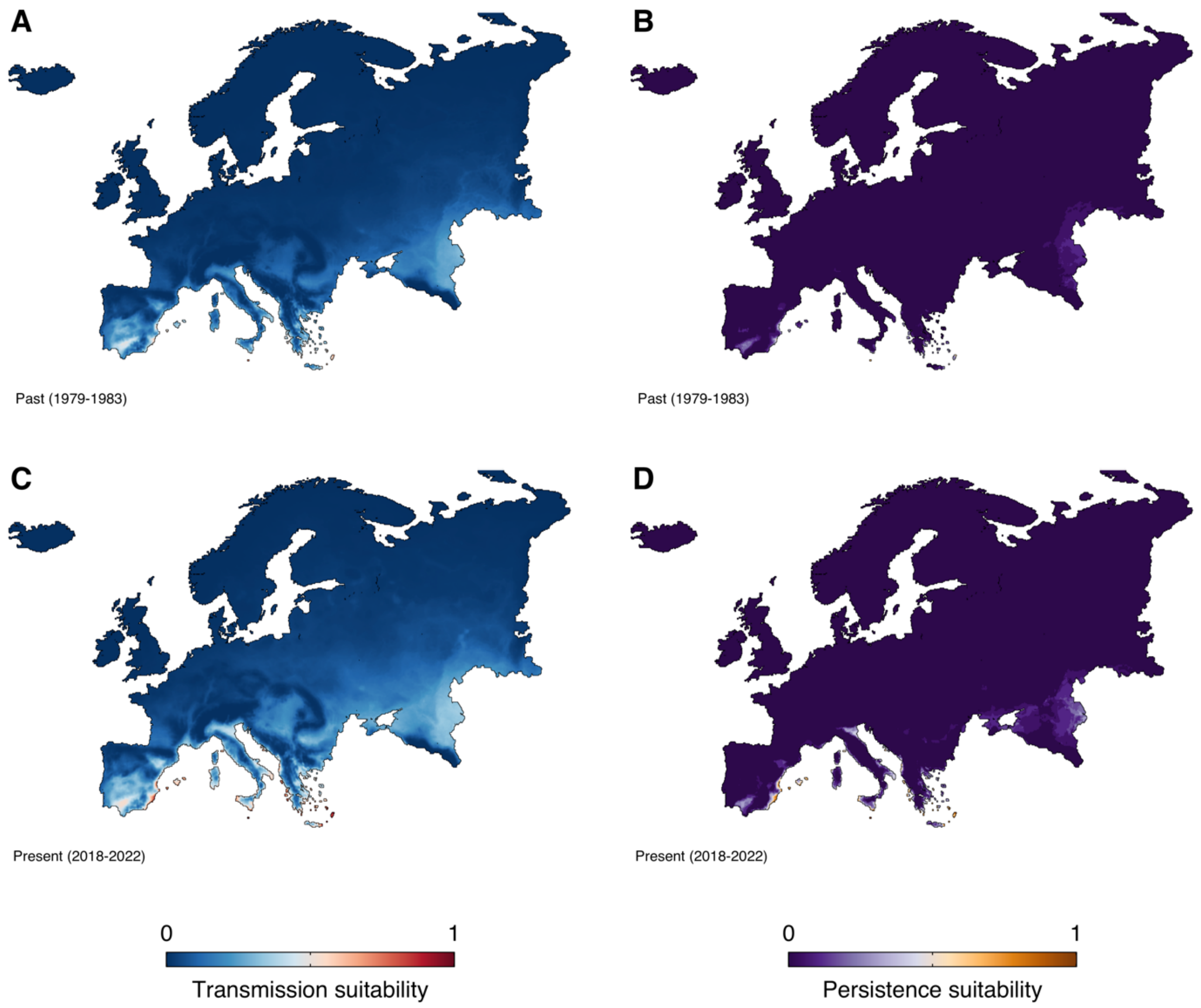


**Fig. S11. Map of past and present DENV transmission and persistence suitability in Europe.** (**A**, **C**). Average transmission suitability for 1979-1983 and 2018-2022 on a normalized scale, respectively. (**B**, **D**) Average persistence suitability for 1979-1983 and 2018-2022 on a normalized scale, respectively.

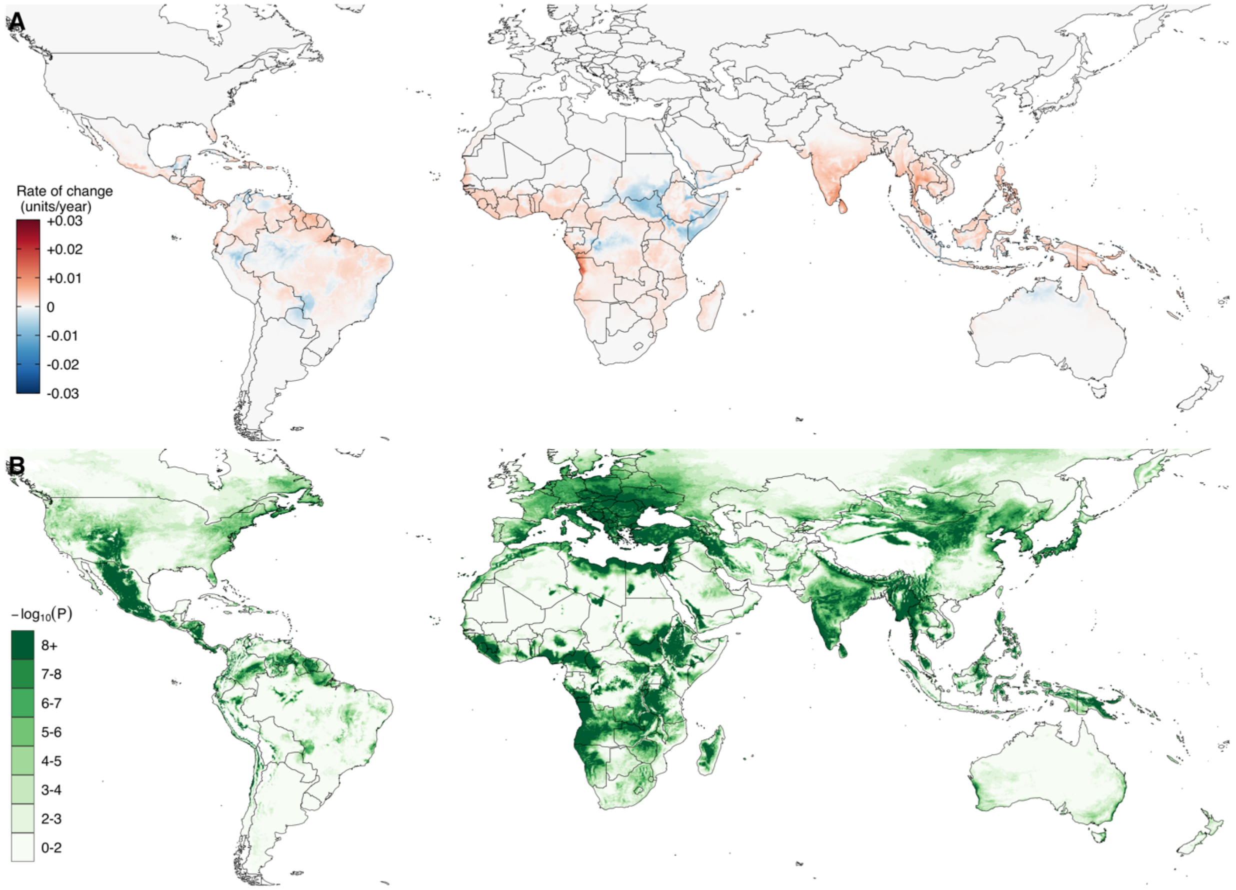


**Fig. S12. Global map of the magnitude and uncertainty of historical climate stress.** (**A**) Estimated yearly change in climate-based dengue transmission suitability (i.e. climate stress) per spatial pixel from 1979-2022 using the seasonal Mann-Kendall (MK) trend test and Sen's slope on monthly Index P time series. (**B**) Associated p-values from the seasonal MK trend test.
